# Exponentially weighted ensemble deep learning for histologic growth pattern classification in lung adenocarcinoma

**DOI:** 10.64898/2026.09.24.26363902

**Authors:** Rajashekar Korutla, Muskan Naresh Jain, Tejas Mohan Karkera, Anne Breggia, Robert Christman, Stephen T. Ryan, Gene Tunik, Saeed Amal

## Abstract

Accurate classification of lung adenocarcinoma growth patterns from hematoxylin and eosin (H&E)-stained whole-slide images is essential for treatment planning and prognosis. We develop and evaluate an exponentially weighted ensemble deep learning approach that combines five architectures (EfficientNet-B3, DeiT3, Swin Transformer, Vision Transformer, ConvNeXt) with weights determined by a softmax transformation of per-model Quadratic Weighted Kappa (QWK) scores. Using 25,545 image patches at 40× magnification from the Dartmouth-Hitchcock dataset, we evaluated performance with stratified 5-fold cross-validation. Individual model performance ranged from Cohen’s kappa *κ* = 0.5563 (ConvNeXt) to *κ* = 0.9561 (EfficientNet-B3). The ensemble achieved 96.52% accuracy and *κ* = 0.9648, exceeding the best individual model by Δ*κ* = +0.0087 with statistical significance (McNemar’s test, *χ*^2^ = 50.84, *p <* 10^*−*12^; bootstrap 95% CI on Δ*κ* [+0.0056, +0.0117]). Pairwise disagreement rates of 16.6–43.2% between architectures indicate that the models capture different aspects of tissue morphology, and the framework automatically down-weights unreliable components, as illustrated by ConvNeXt receiving only 5.7% ensemble weight despite a single-fold training failure. The approach offers a clinically grounded method for ensemble aggregation in digital pathology.

## 1 Introduction

Lung cancer remains the leading cause of cancer-related mortality worldwide, accounting for approximately 1.8 million deaths annually [1, 2]. Among the several types of lung cancer, non-small cell lung cancer (NSCLC) accounts for approximately 80% to 85% of all cases. Adenocarcinoma is the most prevalent histological subtype, representing approximately 40% of all lung cancer diagnoses and roughly 50% of all NSCLC cases [3]. While lung cancer claimed an estimated 127,070 lives in 2023, mortality rates continue to decline due to early detection and treatment advances [2].

Precise detection and categorization of growth patterns in lung adenocarcinomas are essential for directing therapeutic approaches and forecasting patient outcomes. The World Health Organization (WHO) 2015 classification system identifies five primary growth patterns for lung adenocarcinoma: lepidic, acinar, papillary, micropapillary, and solid [4]. These patterns exhibit distinct morphological characteristics: the lepidic pattern shows tumor cells growing along existing alveolar structures; the acinar pattern displays glandular architecture with rounded spaces; the papillary pattern exhibits finger-like projections with fibrovascular cores; the micropapillary pattern contains small papillary tufts floating in alveolar spaces; and the solid pattern demonstrates sheets of tumor cells without glandular architecture [4, 5]. The WHO classification emphasizes the importance of recording minor components, as lung adenocarcinomas frequently display a heterogeneous mixture of several patterns within a single tumor [4].

Identifying these growth patterns through hematoxylin and eosin (H&E)-stained images is crucial because they help distinguish invasive and non-invasive subtypes. For example, accurate identification of the lepidic growth pattern aids in predicting invasive adenocarcinoma subtypes including adenocarcinoma in situ (AIS), minimally invasive adenocarcinoma (MIA), and invasive lepidic predominant adenocarcinoma (iLPA) [5]. However, manual identification of histologic subtypes presents several challenges, including interobserver variability, where different pathologists may assign different diagnoses to the same tissue sample, as well as extended diagnostic timelines with potential misinterpretations due to the laborious nature of the process [4, 6].

Recent developments in deep learning have advanced image analysis methods in medical imaging [7, 8]. Convolutional neural networks (CNNs) enable data-driven extraction of relevant features from histopathological images [7]. However, individual deep learning models have shown limitations in achieving clinical-grade performance for complex histopathological tasks. Previous studies on lung adenocarcinoma classification have reported moderate success, with the best individual model achieving *κ* = 0.525 on the Dartmouth dataset [9], well below the threshold typically associated with high interobserver reliability.

Cohen’s Kappa (*κ*) is a statistical measure of inter-rater agreement that accounts for chance agreement, widely used in pathology to evaluate concordance between automated systems and expert pathologists [10]. The interpretation follows established guidelines: *κ* 0.00–0.20 indicates slight agreement, 0.21–0.40 fair, 0.41–0.60 moderate, 0.61–0.80 substantial, and 0.81–1.00 almost perfect agreement [11]. Values of *κ >* 0.8 indicate “almost perfect agreement” between observers [10, 12]. While this threshold is not pathology-specific, it is widely used in medical research to denote a very high level of interobserver reliability, which may be considered suitable for clinical-grade diagnostic agreement in many applications.

While individual architectures have shown promise, ensemble approaches that combine multiple complementary models remain largely unexplored specifically for lung adenocarcinoma histologic classification. Each neural network architecture captures different aspects of tissue morphology: convolutional networks excel at local texture analysis, while transformer architectures provide superior global context modeling [13, 14]. Recent ensemble studies in medical imaging have shown competitive performance compared to individual models across various cancer classification tasks [15, 16]. However, traditional ensemble methods typically employ simple averaging or accuracy-based weighting [17], which may not align with clinical requirements where inter-rater agreement measured by Cohen’s Kappa is more clinically relevant than raw accuracy metrics.

This work presents an ensemble deep learning approach for automated lung adenocarcinoma histologic subtype classification. We use an exponentially-weighted QWK ensemble strategy that allocates model weights through a softmax transformation of per-model Quadratic Weighted Kappa scores, thereby concentrating ensemble influence on models with the highest clinical agreement while still leveraging diverse architectural perspectives. Our approach combines five architectures (EfficientNet-B3, DeiT3, Swin Transformer, ViT, and ConvNeXt) selected to provide complementary feature extraction capabilities spanning local texture analysis (CNNs) and global context modeling (transformers). The ensemble approach achieves 96.52% accuracy and *κ* = 0.9648 on the standardized Dartmouth dataset, exceeding the best individual model (EfficientNet-B3, *κ* = 0.9561) with *κ >* 0.96 on this benchmark. We provide a complementarity analysis demonstrating that architecturally diverse models capture distinct aspects of tissue morphology, and a computational cost analysis to guide deployment decisions.

### 1.1 Related Work

Recent studies have explored various deep learning approaches for classifying lung adenocarcinoma histological patterns with varying degrees of success. Wei et al. (2019) [9] established the foundational benchmark on the Dartmouth-Hitchcock Medical Center dataset, utilizing a patch classifier with a sliding window technique and reporting a Cohen’s Kappa score of 0.525 on 143 whole-slide images (WSIs), representing moderate agreement with expert pathologists. Sheikh et al. (2022) [18] developed an unsupervised model using stacked autoencoders, achieving 94.60% accuracy on a 5-class classification task using 31 WSIs, though the limited dataset size raises questions about generalizability. DiPalma et al. (2021) [19] employed a resolution-based distillation approach with ResNet, achieving 94.51% accuracy on the 269-slide lung adenocarcinoma dataset; however, they did not report Cohen’s Kappa scores, and acknowledged that their visualization methods still lack full clinical interpretability for pathologists.

Balasubramanian et al. (2024) [16] developed an architectural integration approach in digital pathology by developing an ensemble deep learning framework for breast cancer subtype and invasiveness diagnosis. Utilizing the BACH and BreakHis datasets, their CNN-based ensemble approach (combining VGG16 and ResNet variants) achieved patch-level classification accuracies exceeding 95%. While highly successful for breast tissue, this methodology relied exclusively on convolutional architectures and has not been directly applied to the morphological challenges, such as identifying the five distinct WHO growth patterns, inherent to lung adenocarcinoma classification.

Ali et al. (2023) [20] conducted a comprehensive scoping review of vision transformer applications in lung cancer, analyzing 34 studies and identifying computational complexity and clinical relevance as critical research gaps. More recent works have introduced advanced transformer-based models such as TransPath [21] and HIPT [22] for histopathological image analysis. While these methods report strong performance in various cancer classification tasks, they often require specialized multi-resolution training pipelines with substantial computational overhead. Table 1 summarizes the comparison of existing approaches.

**Table 1:** Comparison of Research Studies.

| Author, Year | Technical Method | Dataset | Classification | Performance |
| --- | --- | --- | --- | --- |
| Wei, 2019 [9] | Patch classifier, sliding window | 143 WSIs, DHMC | 5-class | F1: 90.4%, $\kappa$: 0.525 |
| Sheikh, 2022 [18] | Stacked autoencoders | 31 WSIs, DHMC | 5-class | Acc: 94.60% |
| DiPalma, 2021 [19] | MIL with ResNet | 269 slides, TCGA+DHMC | 5-class | Acc: 94.51% |
| Balasubramanian, 2024 [16] | Ensemble (VGG16+ResNet) | 143 WSIs, DHMC (breast) | Breast subtypes | Acc: 89.8%, $\kappa$: 0.908 |
| Our Study, 2025 | Exp-Weighted QWK Ensemble | 25,545 patches, DHMC | 5-class lung | Acc: 96.52%, $\kappa$: 0.9648 |

## 2 Materials and Methods

Figure 1 provides an overview of the proposed pipeline, from input image preprocessing through parallel feature extraction across the five deep learning backbones to final weighted ensemble aggregation.

**Figure 1:**
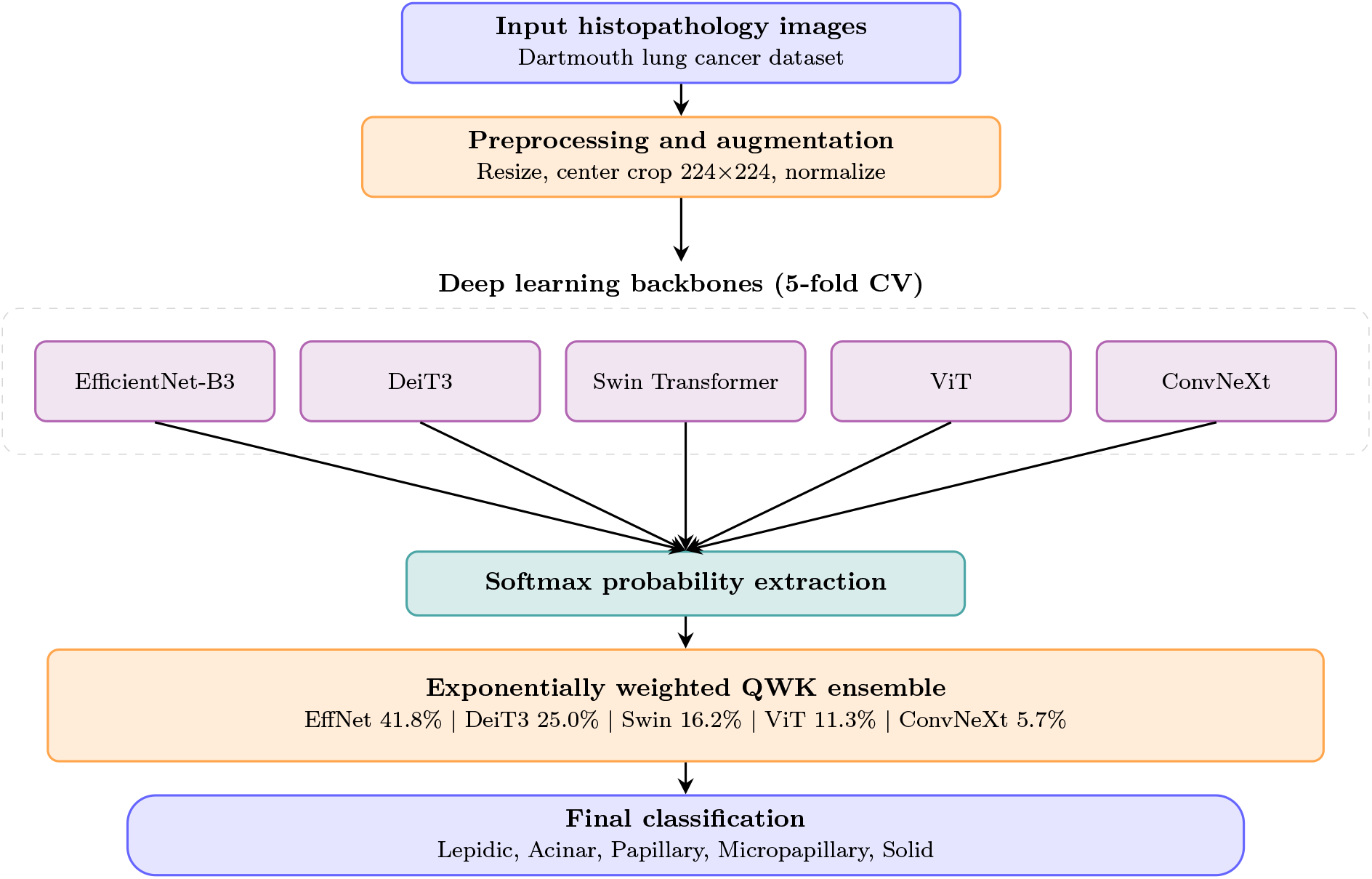
Overview of the proposed exponentially weighted QWK ensemble methodology, showing data preprocessing, parallel feature extraction across the five deep learning backbones, and final weighted aggregation.

### 2.1 Dataset Description

The dataset comprises H&E-stained formalin-fixed paraffin-embedded (FFPE) whole-slide images of lung adenocarcinoma obtained from Dartmouth-Hitchcock Medical Center [23]. A total of 143 WSIs were scanned at 40× magnification, and image patches of 256 × 256 pixels were extracted using the WSITools library [24] with a tissue detection threshold of 0.5 using the Gaussian Naive Bayes (GNB) tissue detector and a minimum tissue area filter of 90% (`patch_filter_by_area=0.90`). This approach targets tissue-rich regions, avoiding background and artifact areas. The extracted patches were categorized into five classes according to the WHO 2015 classification: Lepidic, Acinar, Papillary, Micropapillary, and Solid [4]. Class annotations were provided by expert pathologists at Dartmouth-Hitchcock Medical Center as part of the original dataset curation [9, 23]. The final dataset contains 25,545 patches with 5,109 samples per class. The class balance was established at the patch level through subsampling during dataset curation, distributed across five pre-defined cross-validation folds (1,021 patches per class for Folds 1–4 and 1,025 for Fold 5)

While the overall dataset is balanced at the class level, the number of patients contributing to each class varies substantially: Acinar (51 patients), Papillary (59 patients), Lepidic (19 patients), Micropapillary (9 patients), and Solid (5 patients) (Supplementary Figure S1). This imbalance in patient-level representation is characteristic of the natural prevalence distribution of these histologic subtypes and motivates our use of weighted sampling and focal loss during training.

### 2.2 Data Preprocessing and Augmentation

All extracted patches (256 × 256 pixels) were resized to 224 × 224 pixels to accommodate the fixed input size requirements of all five pretrained architectures, which were originally designed for 224 × 224 ImageNet inputs. Normalization was applied using ImageNet statistics (mean: [0.485, 0.456, 0.406], std: [0.229, 0.224, 0.225]).

A data augmentation strategy was applied uniformly across all classes during training. Augmentations included: random resized cropping (scale range: 0.8–1.0), horizontal flipping (*p* = 0.5), vertical flipping (*p* = 0.3), random rotation (±15 degrees), color jittering (brightness, contrast, saturation, and hue variations within ±10%), Gaussian noise injection (*p* = 0.3), Gaussian blur (*p* = 0.3), and cutout regularization (8 holes, 16 × 16 pixels, *p* = 0.5). The ±15-degree rotation range was selected to minimize border artifacts; at this angle, tissue-free corner regions are negligible relative to the 224 × 224 input size. Class imbalance during training is addressed separately through weighted random sampling. Validation images underwent only resizing and center cropping with normalization. Supplementary Table S1 summarizes all training configuration parameters.

Differentiated batch sizes were used for training (24) and validation (32). Smaller training batches accommodate memory requirements of larger transformer models, while larger validation batches maximize inference throughput. This is consistent with standard practice in deep learning, where training and evaluation batch sizes are independently optimized for gradient stability and inference throughput, respectively.

### 2.3 Model Architectures

Five complementary architectures were selected to provide diverse feature extraction capabilities spanning convolutional and transformer-based paradigms, each initialized with pretrained ImageNet weights for transfer learning:

- **EfficientNet-B3** [25]: A compound scaling architecture (10.7M parameters) that systematically scales network width, depth, and resolution, providing an efficient balance of accuracy and computational cost for histopathological texture analysis.
- **DeiT3** [26]: A data-efficient vision transformer (85.8M parameters) employing a distillation mechanism for effective learning from limited labeled data.
- **Swin Transformer** [27]: A hierarchical vision transformer (86.7M parameters) computing self-attention within local windows and shifting windows between layers, capturing both local cellular structures and global tissue organization.
- **Vision Transformer (ViT)** [13]: The standard vision transformer (85.8M parameters) applying global self-attention to model long-range spatial dependencies in tissue architecture.
- **ConvNeXt** [28]: A modernized convolutional architecture (27.8M parameters) incorporating transformer-inspired design elements while maintaining convolutional efficiency.

These architectures provide complementary feature extraction: EfficientNet-B3 and ConvNeXt deliver local texture analysis through convolutions, while DeiT3, Swin Transformer, and ViT contribute global context through attention mechanisms.

### 2.4 Training and Evaluation Protocol

We employed the stratified 5-fold cross-validation splits provided in the original Dartmouth dataset, where patches are stratified by class label across folds. For each fold, four partitions served as training data and one as the held-out validation set. The AdamW optimizer [29] was used with learning rate 2 × 10^*−*4^ and weight decay 1 × 10^*−*4^. A CosineAnnealingWarmRestarts scheduler (*T*_0_ = 10, *T*_*mult*_ = 2) provided dynamic learning rate adjustment. A combined loss of Focal Loss (*α* = 1, *γ* = 2) at weight 0.7 and Label Smoothing Cross-Entropy (smoothing=0.1) at weight 0.3 was employed. Focal Loss addresses class imbalance by down-weighting well-classified examples [30], while label smoothing [31] prevents overconfident predictions and improves generalization. Weighted random sampling ensured balanced class representation within each batch. Training proceeded for up to 25 epochs with early stopping on validation QWK (patience=7). Gradient clipping (max norm=1.0) ensured stability.

### 2.5 QWK-Weighted Ensemble Strategy

#### Quadratic Weighted Kappa (QWK)

Cohen’s Kappa quantifies inter-rater agreement beyond chance, but treats all disagreements as equally severe. Quadratic Weighted Kappa (QWK) extends Cohen’s Kappa for ordered categorical outcomes by introducing a weighting matrix that penalizes larger ordinal disagreements more heavily than smaller ones [11, 10]. For an *N* -class ordinal problem with confusion matrix *O*, the quadratic weight matrix *W* is defined as:

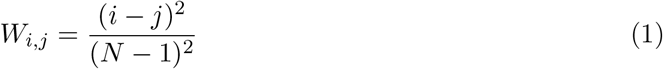

where *W*_*i,j*_ = 0 when predicted class *j* equals true class *i* (perfect agreement), and *W*_*i,j*_ = 1 for the maximally distant misclassification. Let *E* denote the expected agreement matrix under independence (computed from the marginal distributions of the true and predicted labels). The QWK is then:

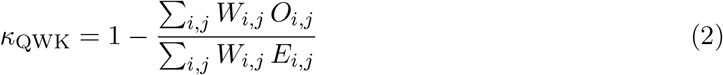

The WHO classification of lung adenocarcinoma growth patterns reflects an approximate clinical ordering by aggressiveness, with lepidic patterns associated with the best prognosis and micropapillary and solid patterns associated with worse outcomes [5]. This makes QWK clinically appropriate for evaluating classification performance on this task: misclassifications between distantly related subtypes (e.g., lepidic vs. solid) carry greater clinical consequence than those between adjacent subtypes (e.g., acinar vs. papillary), and QWK reflects this through its quadratic penalty structure.

#### Exponentially-Weighted QWK Ensemble

The central methodological contribution of this work is the exponentially-weighted QWK ensemble strategy. Let *M* = {*m*_1_, *m*_2_, …, *m*_5_} denote the trained models. For each model *m*_*i*_, the average QWK score across *K* = 5 folds is:

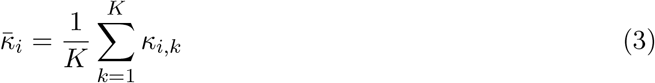

We initially considered linear weighting (weights proportional to 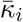), but found that linear weighting underperforms the strongest individual model when the spread of per-model performance is wide. To better concentrate ensemble influence on the highest-performing components while still benefiting from architectural diversity, we adopt a softmax-based exponential weighting:

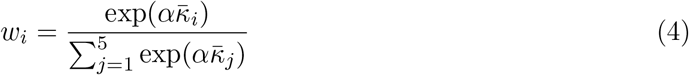

where *α* ≥ 0 is a temperature hyperparameter that controls how aggressively weight is concentrated on top performers. When *α* = 0, all models receive equal weight (simple averaging); as *α* → ∞, the ensemble degenerates to selecting the single best model. We use *α* = 5, which balances concentration on high-performing models with meaningful contribution from all five components. The final ensemble prediction combines weighted softmax probabilities:

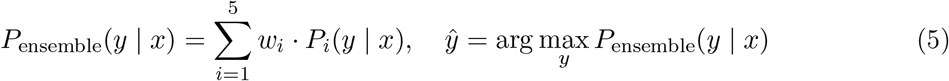

The computed weights with *α* = 5 were: EfficientNet-B3 (41.8%), DeiT3 (25.0%), Swin Transformer (16.2%), ViT (11.3%), and ConvNeXt (5.7%). Notably, ConvNeXt’s low average QWK (0.5563) automatically results in a small ensemble weight (5.7%), so its catastrophic Fold 1 failure has minimal impact on the ensemble’s aggregate performance. Test-time augmentation (TTA) [32] with four transformation variants (original, horizontal flip, vertical flip, rescaled center crop) was applied during inference to reduce prediction variance.

### 2.6 Model Interpretability via Grad-CAM

Gradient-weighted Class Activation Mapping (Grad-CAM) [33] was applied to EfficientNet-B3, the highest-weighted ensemble component (mean *κ* = 0.9561, weight = 41.8%). We targeted the fourth convolutional block (14 × 14 spatial resolution) for high-quality heatmaps.

### 2.7 Statistical Analysis

To assess whether the ensemble’s improvement over individual architectures is statistically significant, we used two complementary tests at the patch level. First, we performed McNemar’s test [10] with continuity correction on the 2 × 2 contingency table of paired correct/incorrect predictions across all *n* = 25,545 patches. McNemar’s test evaluates, for a given pair of classifiers, whether they have equal error rates by examining only the discordant cases (where one classifier is correct and the other wrong) on the same set of samples. We applied the test independently to each of the five pairwise comparisons (ensemble vs. each individual architecture); each test reduces multi-class predictions to a binary correct/incorrect outcome per patch, so the 5-class structure of the underlying task does not affect the test’s validity. All tests were two-tailed with significance threshold *α* = 0.05. Second, we computed bootstrap 95% confidence intervals for the difference in QWK (Δ*κ*) between the ensemble and each individual model using 1,000 resamples of the patch-level predictions with replacement. Confidence intervals that exclude zero indicate a statistically meaningful difference. All performance values reported as mean ± standard deviation across the five folds use the population standard deviation; the notation “±” indicates standard deviation throughout.

## 3 Results

### 3.1 Individual Model Performance

Table 2 presents validation performance across all five folds. EfficientNet-B3 showed the strongest individual performance (95.90% ± 2.23% accuracy, *κ* = 0.9561 ± 0.0261). DeiT3 achieved 84.48% ± 2.99% accuracy (*κ* = 0.8528 ± 0.0275). Swin Transformer showed moderate performance (76.33% ± 5.67%, *κ* = 0.7664 ± 0.0683). ViT achieved 68.78% ± 7.00% (*κ* = 0.6948 ± 0.0789). ConvNeXt showed the most variable performance (58.11% ± 20.09%, *κ* = 0.5563 ± 0.2844), including a complete training failure in Fold 1 (*κ* = 0.0000, accuracy = 20.00%, equivalent to random guessing for a 5-class problem). We retained ConvNeXt as part of the ensemble despite its instability to show how the exponentially-weighted QWK framework handles such failures, automatically assigning low weights to unreliable components.

**Table 2:**
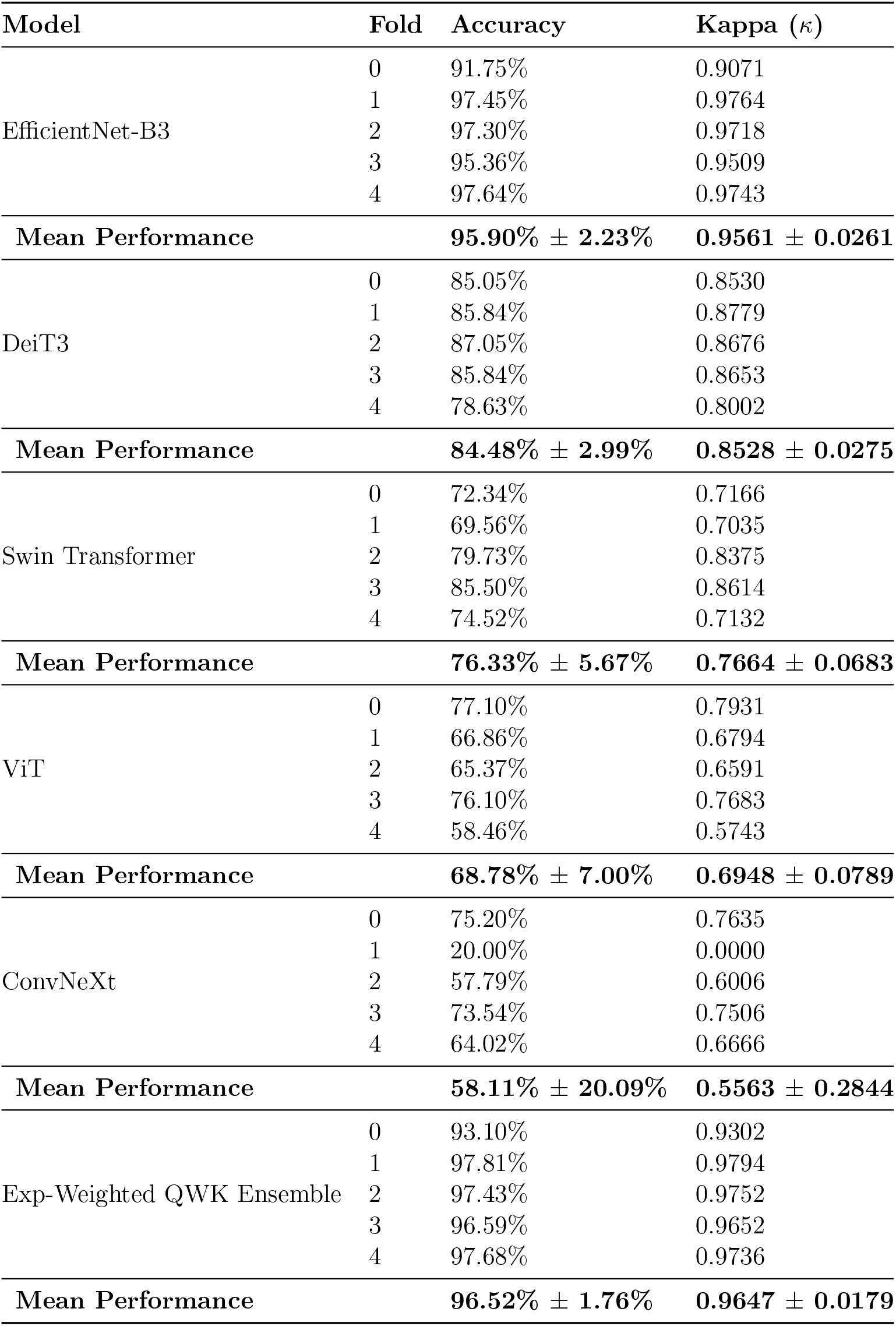
Validation Performance Metrics Across 5-Fold Cross-Validation.

### 3.2 Ensemble Performance and Weighting Comparison

The exponentially-weighted QWK ensemble achieved 96.52% ± 1.76% accuracy and *κ* = 0.9647 ± 0.0179 across the five cross-validation folds (aggregate *κ* = 0.9648 on the pooled predictions of *n* = 25,545 patches), exceeding the best individual model (EfficientNet-B3, *κ* = 0.9561) by Δ*κ* = +0.0087. The ensemble outperformed EfficientNet-B3 in 4 of 5 individual folds (Folds 0, 1, 2, and 3). To assess statistical significance, we applied McNemar’s test on patch-level predictions: out of 491 patches where the two classifiers disagreed, the ensemble was correct on 325 and EfficientNet-B3 on 166 (net +159 in favor of the ensemble), yielding *χ*^2^ = 50.84 and *p* = 1.0 × 10^*−*12^. Bootstrap 95% confidence intervals on Δ*κ* (1,000 resamples of the patch-level predictions) yielded [+0.0056, +0.0117], excluding zero and supporting the improvement. McNemar’s tests against the other four architectures (DeiT3, Swin Transformer, ViT, and ConvNeXt) all yielded *p <* 10^*−*300^ in favor of the ensemble (Table 3).

**Table 3:** McNemar’s Test: Ensemble vs. Each Individual Architecture (*n* = 25,545 patches)

| Comparison | Both Correct | Ensemble Wins | Individual Wins | Both Wrong | $\chi^2$ | $p$-value |
| --- | --- | --- | --- | --- | --- | --- |
| Ensemble vs. EfficientNet-B3 | 24,332 | 325 | 166 | 722 | 50.84 | $1.0 \times 10^{-12}$ |
| Ensemble vs. DeiT3 | 21,325 | 3,332 | 255 | 633 | 2,637.80 | $< 10^{-300}$ |
| Ensemble vs. Swin Transformer | 19,275 | 5,382 | 223 | 665 | 4,746.65 | $< 10^{-300}$ |
| Ensemble vs. ViT | 17,335 | 7,322 | 232 | 656 | 6,652.62 | $< 10^{-300}$ |
| Ensemble vs. ConvNeXt | 14,637 | 10,020 | 208 | 680 | 9,411.00 | $< 10^{-300}$ |

To examine the effect of the weighting scheme, we compared several variants of the QWK-weighting framework, all using the same five trained models and the same predictions, differing only in how per-model weights are derived from the average QWK scores. Supplementary Table S2 reports the results.

The exponential weighting with *α* = 5 provides the best balance: it concentrates roughly 42% of the ensemble’s influence on EfficientNet-B3 (the strongest individual model) while still allocating 58% across the four other architectures, allowing the ensemble to benefit from architectural diversity. Increasing *α* further (e.g., *α* = 10) over-concentrates on EfficientNet-B3 and erodes the diversity benefit, while linear weighting under-weights the strongest model and underperforms it.

Per-class accuracy of the exponentially-weighted ensemble: Lepidic (95.87%), Acinar (96.28%), Papillary (95.05%), Micropapillary (97.79%), and Solid (97.63%). The ensemble achieves both higher overall agreement and more uniform per-class performance than any individual model.

### 3.3 Computational Cost Analysis

Supplementary Table S3 presents computational requirements. EfficientNet-B3 offers the strongest accuracy-to-cost ratio with only 10.7M parameters and 626 images/second throughput. The full ensemble requires 296.8M total parameters and processes approximately 79 images/second, a 7.9× reduction in throughput. This computational overhead must be weighed against the ensemble’s *κ* improvement of +0.0087 over EfficientNet-B3 alone. The ensemble’s value lies in three aspects: (1) *higher aggregate clinical agreement* : *κ* = 0.9648 vs. *κ* = 0.9561, a measurable improvement that may be relevant for diagnostic decision support; (2) *robustness to model failure*: ConvNeXt’s catastrophic collapse in Fold 1 (*κ* = 0.000) is automatically de-emphasized by the exponential weighting, so individual architecture failures have minimal impact on the ensemble; and (3) *extensibility*: the QWK-weighting framework can incorporate future models without retraining existing components. For high-throughput screening where speed is paramount, EfficientNet-B3 alone is a strong single-model option. For settings prioritizing aggregate agreement and robustness, the ensemble’s 79 img/s enables analysis of a typical WSI (1,000–2,000 patches) in under 30 seconds, which remains practical for clinical workflows.

### 3.4 Model Complementarity Analysis

Supplementary Table S4 presents pairwise disagreement rates between all model pairs. Disagreement ranges from 16.6% (EfficientNet-B3 vs. DeiT3, the most similar pair) to 43.2% (Swin Transformer vs. ConvNeXt, the most diverse pair), confirming that each architecture captures distinct aspects of tissue morphology. Per-class accuracy analysis (Table 4) reveals complementary strengths: EfficientNet-B3 achieves uniformly high accuracy (94.1–97.2%), while ConvNeXt shows strong Micropapillary performance (83.2%) but struggles with Papillary (45.8%), and ViT shows stronger Solid classification (83.5%) but weaker Acinar performance (59.4%). The ensemble achieves high and more uniform per-class performance (95.1–97.8%) while integrating these complementary architectural strengths, with notable gains over EfficientNet-B3 alone on Solid (+0.6 percentage points) and Micropapillary (+0.6 pp) classes.

**Table 4:** Per-Class Accuracy by Architecture (%)

| Model | Lepidic | Acinar | Papillary | Micropap. | Solid |
| --- | --- | --- | --- | --- | --- |
| EfficientNet-B3 | 95.3 | 95.8 | 94.1 | 97.2 | 97.0 |
| DeiT3 | 83.6 | 84.3 | 75.5 | 89.3 | 89.8 |
| Swin Transformer | 77.1 | 71.7 | 61.9 | 88.5 | 82.5 |
| ViT | 65.6 | 59.4 | 65.0 | 70.4 | 83.5 |
| ConvNeXt | 50.0 | 46.1 | 45.8 | 83.2 | 65.4 |
| **Ensemble** | **95.9** | **96.3** | **95.1** | **97.8** | **97.6** |

### 3.5 Confusion Matrix

The ensemble confusion matrix (Table 5) shows strong diagonal dominance. The most common misclassifications involve Papillary confused with Acinar (119 cases) and Lepidic with Papillary (92 cases), consistent with known morphological similarities between these subtypes.

**Table 5:** Ensemble Confusion Matrix.

| True \ Pred | Lepidic | Acinar | Papillary | Micropap. | Solid |
| --- | --- | --- | --- | --- | --- |
| Lepidic | 4898 | 63 | 92 | 35 | 21 |
| Acinar | 20 | 4919 | 86 | 39 | 45 |
| Papillary | 61 | 119 | 4856 | 38 | 35 |
| Micropap. | 12 | 51 | 26 | 4996 | 24 |
| Solid | 41 | 19 | 18 | 43 | 4988 |

### 3.6 Model Interpretability Results

Grad-CAM visualizations from EfficientNet-B3 (Figure 2) show attention patterns on tissue regions: lepidic patterns show attention on alveolar walls; acinar on glandular spaces; papillary on fibrovascular projections; micropapillary on floating tufts; and solid on dense cellular sheets.

**Figure 2:**
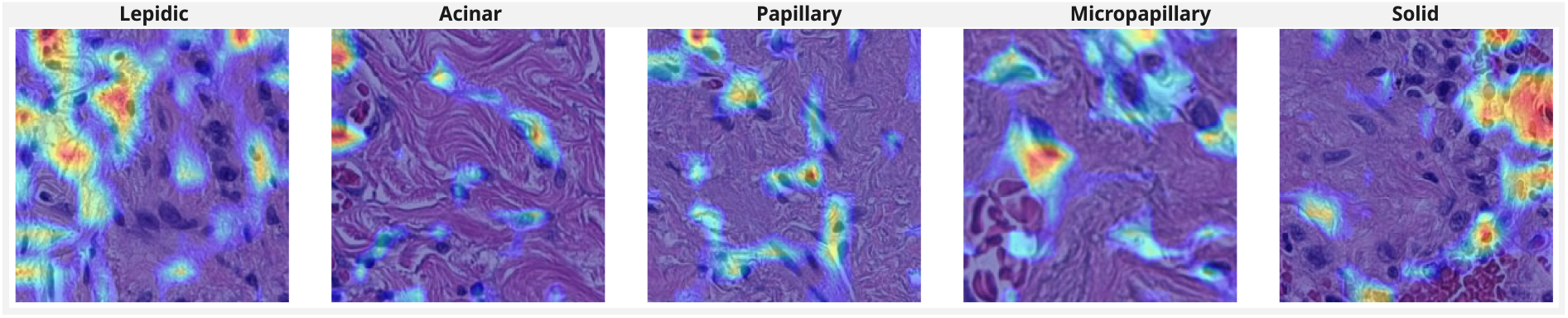
Grad-CAM interpretability visualization on the EfficientNet-B3 backbone, demonstrating accurate morphological targeting across the five adenocarcinoma subtypes.

## 4 Discussion

This study evaluates an exponentially-weighted QWK ensemble approach for lung adenocarcinoma histologic pattern classification, achieving *κ* = 0.9648 on the standardized Dartmouth dataset, an improvement over the prior benchmark of *κ* = 0.525 reported by Wei et al. (2019) [9]. EfficientNet-B3 alone achieved *κ* = 0.9561, consistent with the training methodology, including the combined Focal Loss and Label Smoothing objective [30, 31], weighted sampling, and test-time augmentation [32]. The exponentially-weighted ensemble exceeds this individual baseline by Δ*κ* = +0.0087, supported by McNemar’s test on patch-level predictions (*χ*^2^ = 50.84, *p <* 10^*−*12^) and by bootstrap 95% confidence intervals on Δ*κ* ([+0.0056, +0.0117]) that exclude zero. While the absolute magnitude of the improvement is modest, its consistency across the 25,545 patches suggests that aggregation of complementary architectures provides a measurable improvement in clinical agreement over the strongest single model.

The choice of weighting function is critical. Linear QWK weighting, in which each model’s weight is proportional to its average QWK, underperforms the strongest individual model when the spread of per-model performance is wide, because it dilutes a strong learner with several weaker ones. Our results in Supplementary Table S2 make this trade-off explicit: linear weighting yields *κ* = 0.9523 (Δ*κ* = −0.0038 vs. EfficientNet-B3), while exponential weighting with *α* = 5 yields *κ* = 0.9648 (Δ*κ* = +0.0087). The exponential transformation *w*_*i*_ ∝ exp 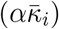 is a well-established mechanism for concentrating weight on top-performing components (it is the same softmax mechanism used in attention and in many ensemble selection schemes), and the temperature *α* provides a single, interpretable knob for trading off concentration against diversity. Selecting *α* = 5 allocates 41.8% of the ensemble’s influence to EfficientNet-B3 while still leveraging the four remaining architectures, capturing a meaningful fraction of the diversity reflected in the 16.6–43.2% pairwise disagreement rates.

ConvNeXt’s catastrophic failure in Fold 1 (20.0% accuracy, *κ* = 0.000) likely reflects training instability specific to that fold’s training trajectory, possibly related to the architecture’s known sensitivity to learning rate and initialization. Importantly, the exponentially-weighted QWK framework handled this gracefully: ConvNeXt’s low overall mean QWK (0.5563) translates to a small ensemble weight of 5.7%, so its single-fold collapse has minimal impact on aggregate ensemble performance. This automatic down-weighting of unreliable components is a key practical advantage of the framework over equal-weighted averaging or accuracy-based weighting, particularly for deployment scenarios involving heterogeneous data sources.

The ensemble requires approximately 7.9× more computation than EfficientNet-B3 alone. The choice between ensemble and EfficientNet-B3 alone therefore depends on deployment context. For high-throughput screening where speed is paramount, EfficientNet-B3 (626 img/s, *κ* = 0.9561) is preferred. For settings prioritizing aggregate agreement and robustness against single-model failures, the ensemble (79 img/s, *κ* = 0.9648) provides a measurable accuracy gain at a still-practical processing rate.

### 4.1 Comparison with State-of-the-Art

Recent methods including TransPath [21] and HIPT [22] leverage multi-scale hierarchical learning for histopathological image analysis. These approaches often require specialized multi-resolution training pipelines with substantial computational overhead and custom architectures. In contrast, our approach achieves strong performance (*κ >* 0.96) through intelligent combination of standard, readily available architectures. The QWK-weighting framework offers practical advantages through its modular design: individual components can be independently updated, replaced, or augmented as new architectures emerge, without requiring retraining of the entire system. The Balasubramanian et al. (2024) [16] ensemble study on breast cancer achieved *κ* = 0.908 using CNN-only architectures on the same institutional data source. Our study extends this ensemble paradigm to lung adenocarcinoma with a broader architectural repertoire that includes both CNNs and transformers, and introduces the exponentially-weighted QWK scheme as an alternative to equal-weighted or accuracy-based averaging [15, 17].

### 4.2 Limitations

Several limitations should be acknowledged. First, evaluation is conducted on a single institutional dataset; multi-institutional testing with diverse patient populations, staining protocols, and scanning equipment is necessary to establish broader generalizability. Second, the patient-level class distribution is highly skewed, with only 5 patients contributing all Solid patches and 9 patients contributing all Micropapillary patches. Furthermore, within these under-represented classes, patches are dominated by a small number of patients: in the Solid class, a single patient contributes 2,370 of 5,109 patches (46%); in Micropapillary, one patient contributes 2,104 patches (41%); and in Lepidic, one patient contributes 1,978 patches (39%). Combined with the patch-level fold assignment provided in the original Dartmouth dataset, which does not enforce patient-level separation across folds, the high per-class metrics for these three subtypes may partly reflect intra-patient tissue consistency rather than generalization to unseen patients. Patient-level cross-validation on a larger and more demographically diverse cohort is necessary to disentangle these effects and is a top priority for future work. Third, our approach operates at the patch level without slide-level aggregation strategies; clinical deployment would benefit from integrating patch-level predictions into slide-level diagnostic decisions. fourth, the ensemble’s 296.8M total parameters may present deployment challenges in resource-constrained environments. fifth, the system does not integrate molecular markers, immunohistochemistry, or clinical context, all of which inform pathologist decision-making in practice. Finally, scanner variability across institutions and demographic variations in tissue characteristics have not been explicitly evaluated and represent important directions for future investigation.

### 4.3 Ethical Considerations

Prospective clinical implementation studies should incorporate institutional review board approval, informed consent protocols, and assessment of the system’s performance across diverse demographic populations. The system should be viewed as a diagnostic support tool to augment, not replace, expert pathologist judgment.

### 4.4 Future Directions

Future research includes: (1) multi-institutional validation across diverse patient populations and scanner platforms; (2) evaluation of emerging architectures and edge deployment optimizations; (3) multimodal integration with genomic and clinical data; (4) prospective clinical studies with appropriate ethical review; (5) ensemble-aware interpretability methods that account for contributions from all five architectures rather than focusing solely on the highest-weighted component; (6) systematic demographic subgroup analysis to assess fairness and equity; and (7) principled methods for selecting the temperature parameter *α* in the exponential weighting scheme, including nested cross-validation and Bayesian approaches.

## 5 Conclusion

This study shows that an exponentially weighted ensemble of five deep learning architectures, with weights derived from a softmax transformation of per-model Quadratic Weighted Kappa scores, achieves *κ* = 0.9648 on the Dartmouth lung adenocarcinoma dataset and exceeds the strongest individual model (EfficientNet-B3, *κ* = 0.9561) with statistical significance (McNemar’s test, *p <* 10^*−*12^; bootstrap 95% CI on Δ*κ* [+0.0056, +0.0117]). The QWK-based weighting offers a clinically grounded alternative to accuracy-weighted or equal-weighted aggregation in digital pathology, and the exponential temperature parameter provides a tunable balance between concentrating influence on the strongest model and leveraging architectural diversity. The framework is modular and can incorporate new architectures without retraining existing components.

## Author Contributions (CRediT)

Conceptualization, S.A.; Methodology, R.K., M.N.J., T.M.K., and S.A.; Software, R.K., M.N.J., and T.M.K.; Validation, R.K., A.B.; Formal Analysis, R.K., M.N.J., and S.A.; Investigation, R.K., R.C., and S.T.R.; Resources, S.A.; Writing–Original Draft, R.K. and M.N.J.; Writing– Review & Editing, R.K., G.T., and S.A.; Visualization, R.K. and M.N.J.; Supervision, S.A.; Project Administration, S.A. All authors have read and agreed to the published version of the manuscript.

## Data Availability Statement

The dataset is publicly available at: https://bmirds.github.io/LungCancer/

## Ethics Approval

Not applicable. This study uses a publicly available, de-identified dataset.

## Acknowledgments

We thank the Roux Institute, the Institute for Experiential AI, Dr. Bilal Ahmad from Maine Medical Center, and the Alfond Foundation for their support. AI-assisted tools (Claude, Anthropic) were used to support language editing and code review during manuscript preparation. All scientific content, experimental design, and results analysis were performed by the authors. No AI tools were used to create or modify any images in this manuscript.

